# Beliefs and Behaviours related to Antibiotic Use and Antimicrobial Resistance in Poland: General Population Survey

**DOI:** 10.64898/2026.09.15.26363114

**Authors:** Iga Palacz-Poborczyk, Julia Kuzminska, Aleksandra Luszczynska, Aleksandra J. Borek

## Abstract

**Background:** Overuse of antibiotics accelerates antimicrobial resistance (AMR), threatening public health. Poland faces challenges with high antibiotic use; therefore, understanding people’s antibiotic-related behaviours and beliefs would help identify targets for interventions. This study aims to identify sociodemographic and psychosocial correlates of antibiotic use among a representative sample of adults in Poland.

**Methods:** The study employed an exploratory survey design. Adults in Poland (N=1,000) were randomly sampled, ensuring the representativeness of the Polish general population. Data were collected using a pretested questionnaire, administered through computer-assisted telephone interviews. Quantitative data were analysed descriptively and using a logistic regression. Free-text data were analysed using content analysis.

**Results:** 31.6% of respondents used oral antibiotics in the preceding year, most commonly for respiratory tract infections (RTIs). Higher age, higher household income level, and education level were identified as predictors of increased antibiotic use, with increasing age being the most prominent factor. The belief that antibiotics accelerate recovery from RTIs and expectations for antibiotic prescriptions were found to be associated with greater antibiotic use. Encouragingly, 92.9% of respondents reported trusting their doctors, 51.5% that doctors explained why antibiotics were unnecessary for particular infections, and 57.1% were willing to engage in such discussions. While 76.5% of respondents reported awareness of antibiotic resistance, descriptions of the concept were often flawed.

**Conclusions:** Key areas of focus for future interventions include addressing attitudes and behaviours of middle-aged (35-65) and older (66+ years old) adults, managing patient expectations regarding recovery and prescriptions, and correcting fundamental misconceptions about AMR.

## Introduction

Antimicrobial Resistance (AMR) is a critical global public health challenge, threatening our ability to treat infections (1). This escalating crisis undermines modern medicine, making common procedures risky and increasing the burden of infectious diseases worldwide (2). The primary cause of AMR is the widespread misuse and overuse of antibiotics (3). Practices, such as unnecessary prescriptions or self-medication, accelerate the development of resistant microorganisms, reducing drug efficacy (4).

Leading international organisations have maintained commitment to combating AMR through the implementation of coordinated strategies. The WHO Global Action Plan on AMR (1) focuses on five objectives: improving awareness, strengthening surveillance, reducing infection incidence, optimising antimicrobial use, and ensuring sustainable investment. The European One Health Action Plan against AMR (5) focuses on making the European Union (EU) a best-practice region, driving efforts to reduce antimicrobial consumption and improve stewardship across all member states. However, except from temporary decline observed during the COVID-19 pandemic, EU countries are struggling to reduce antibiotic consumption and reach the established reduction targets (6).

Poland is a particularly illustrative case of the challenges to reduce antibiotic consumption, with increased antibiotic consumption in recent years, despite having some of the most rigid reduction goals within the EU (expected antibiotic consumption reduction by 27% until 2030) (6). Evidence in Poland consistently indicated a lack of sufficient knowledge regarding prudent antibiotic use (7). This was accompanied by misconceptions about antibiotic effectiveness for viral infections (8,9). Furthermore, there were concerns regarding patient dissatisfaction with the guidance from healthcare professionals (9) or taking antibiotics without professional consultation (10). Additionally, the concept of AMR was found to be generally unfamiliar (11). At the same time, Poland’s healthcare expenditure and life expectancy both fall below the EU average, while behavioural risk factors for disease are more prevalent than in other regional counterparts (12).

Antibiotic use is not a solely clinical issue, but it is influenced by a range of psychosocial factors (13), such as personal experiences of antibiotic use or beliefs related to antibiotic treatment. Despite a growing recognition of the importance of psychosocial factors in influencing health behaviours, including antibiotic use, significant research gaps persist (1,14). Research has provided initial insights into how *sociodemographic* factors influence antibiotic use (15–17). However, a comprehensive understanding of *psychosocial factors* remains elusive, particularly within different cultural and healthcare contexts.

An evidence inspired, comprehensive, theoretical framework can help identify important behavioural determinants and, then, guide the development of targeted interventions. For example, the Capability, Opportunity, Motivation, and Behaviour (COM-B) model (18) classifies the types of behavioural determinants and can be used to frame important determinants of antibiotic-related behaviours. It has three intervention-relevant domains: (1) Capability, which encompasses the psychological and physical ability to perform the behaviour, such as knowledge and understanding of AMR; (2) Opportunity, which covers external factors that enable or constrain the behaviour, such as access to antibiotic sources and social influences/encouragement from others; (3) Motivation that includes internal processes that direct behaviour, such as beliefs related to antibiotic effectiveness, expectations from prescribers, and concerns related to AMR or antibiotic use itself. The effectiveness and sustainability of behaviour change interventions would be severely impeded if any of the components remains unaddressed.

While previous research has often focused on knowledge assessments (8,19), evidence on the other psychosocial determinants (e.g. motivation, opportunity) of antibiotic use in Poland remains sparse. Moreover, studies have often concentrated on small or local populations (9,20). This creates a critical gap, as a current, nuanced understanding of the general public’s and patients’ beliefs and experiences in Poland is essential for developing effective, national interventions to prevent AMR. Thus, this study aimed to:

1. Describe self-reported behaviours, experiences, and beliefs related to antibiotic use and AMR among adults in Poland.
2. Explore potential associations between sociodemographic characteristics, self-reported behaviours, experiences, and beliefs related to antibiotic use.

## Materials and Methods

This was an observational cross-sectional study among a representative sample of adults in Poland, using an exploratory survey design. It was preregistered in the Open Science Framework repository (21) and is reported according to the STROBE checklist (22) (Supplementary Materials). This study is a part of a larger programme of research (23), aimed at identifying and addressing reasons for high antibiotic consumption in primary care in Poland to contribute to the development of a future intervention.

### Participants

The survey was conducted with members of the general population in Poland. To be included in the study, individuals needed to be at least 18 years old, have a mobile phone number registered in Poland (to enable recruitment), communicate in the Polish language, provide consent and engage in the research. We surveyed 1,000 respondents. This allowed for a maximum margin of error of ±3.1% at a 95% confidence level, a widely accepted standard for representative public opinion surveys.

Respondents were recruited randomly, using mobile phone registers available to a professional public opinion polling organisation. Mobile phone coverage is high in Poland, with 97% of people (aged 15+) using mobile phones (24). Respondents were sampled using stratified random sampling to ensure sample representativeness based on: sex, age category, voivodeship (i.e. county/province), settlement size, and level of education.

### Data Collection

The questionnaire aimed to address the key topics (domains) from the systematic review of 22 public surveys of knowledge and awareness of antibiotic use and AMR from various countries (25). We have also assessed the existing public/patient surveys in Poland (7–11,20) through a mapping review of Polish literature related to antibiotic use and AMR (26). We used these sources to develop a comprehensive list of potential questions, and then selected key questions focusing on less-explored topics to expand the current evidence base.

The questionnaire was pre-tested (N=14), using a think-aloud approach, to check the clarity of wording and that the questions were understood as intended. After three rounds of pre-testing and revisions, unclear or redundant questions were eliminated. The questionnaire was then pilot-tested (N=15) by the polling company, leading to final, minor amendments.

The research protocol was reviewed and approved by the institutional ethics committee. Respondents were briefed on the topic, purpose, length, and anonymity of the survey and gave verbal consent to participate. The anonymous survey collected no personal identifiers, and data were reported in aggregate. Participants could withdraw at any point without consequence, with withdrawn data excluded from the study.

The final questionnaire (Supplementary Materials) consisted of 22 questions (21 structured questions, 1 free-text question) organised in the following sections: (1) experiences with antibiotic treatment (including: antibiotic use, sources of antibiotics, experiences of side effects, self-management, and diagnostic testing); (2) opinions on antibiotics (including: beliefs related to antibiotic efficacy, expected recovery, and side effects; perceived social norms and expectations, expectations for prescriptions and doctor-patient relationship); (3) opinions on AMR (including: concern about AMR, awareness of the AMR term and its understanding). The questionnaire was administered through computer-assisted telephone interviews, lasting on average 12 (7–21) minutes, between 15 and 29 April 2025 – following Poland’s most intense influenza season in years (27).

### Data Analysis

We performed descriptive statistical analysis to characterise the sample and response distributions. Preliminary chi-squared tests identified potential predictors for the subsequent multinomial logistic regression, which modelled the probability of specific categorical outcomes. Age categories were grouped according to American Psychology Association categories of adulthood age classification (28). Settlement size and income level categories were grouped according to classification commonly used in Poland (29,30). The dataset contained no missing values. Responses: “don’t know” and “don’t remember” were excluded from inferential analyses but retained for descriptive statistics. We adopted an alpha level of p≤0.05 for all statistical inference procedures. We used IBM SPSS Statistics (version: 29.0.2.0).

We coded free-text comments on other reasons for which antibiotics were used into overarching categories. Free-text associations with the term antibiotic resistance were analysed using qualitative content analysis. Three researchers independently and inductively coded 10% of the responses, developing a preliminary codebook. After coding another 10% and refining the codebook, one researcher coded the remaining responses, with a second researcher checking all coding for consistency. Any discrepancies were resolved by the team. The coded responses were quantified and synthesised narratively.

## Results

Sample (N=1,000) sociodemographic characteristics, as compared to the Polish population, are reported in Supplementary Materials. Overall, 18,017 individuals were approached: 3,366 declined to participate, 43 discontinued the surveys (data were excluded), and the remaining did not answer or did not align with the sampling frame. Detailed results tables are available in Supplementary Materials.

### Antibiotic use

Overall, 67.8% of respondents reported not taking antibiotics orally and 84.3% not using antibiotic ointments in the preceding year. Single courses of oral antibiotics were the most common, with respiratory tract infections (RTIs) being the primary cause (Fig. 1a).

**Figure 1.**
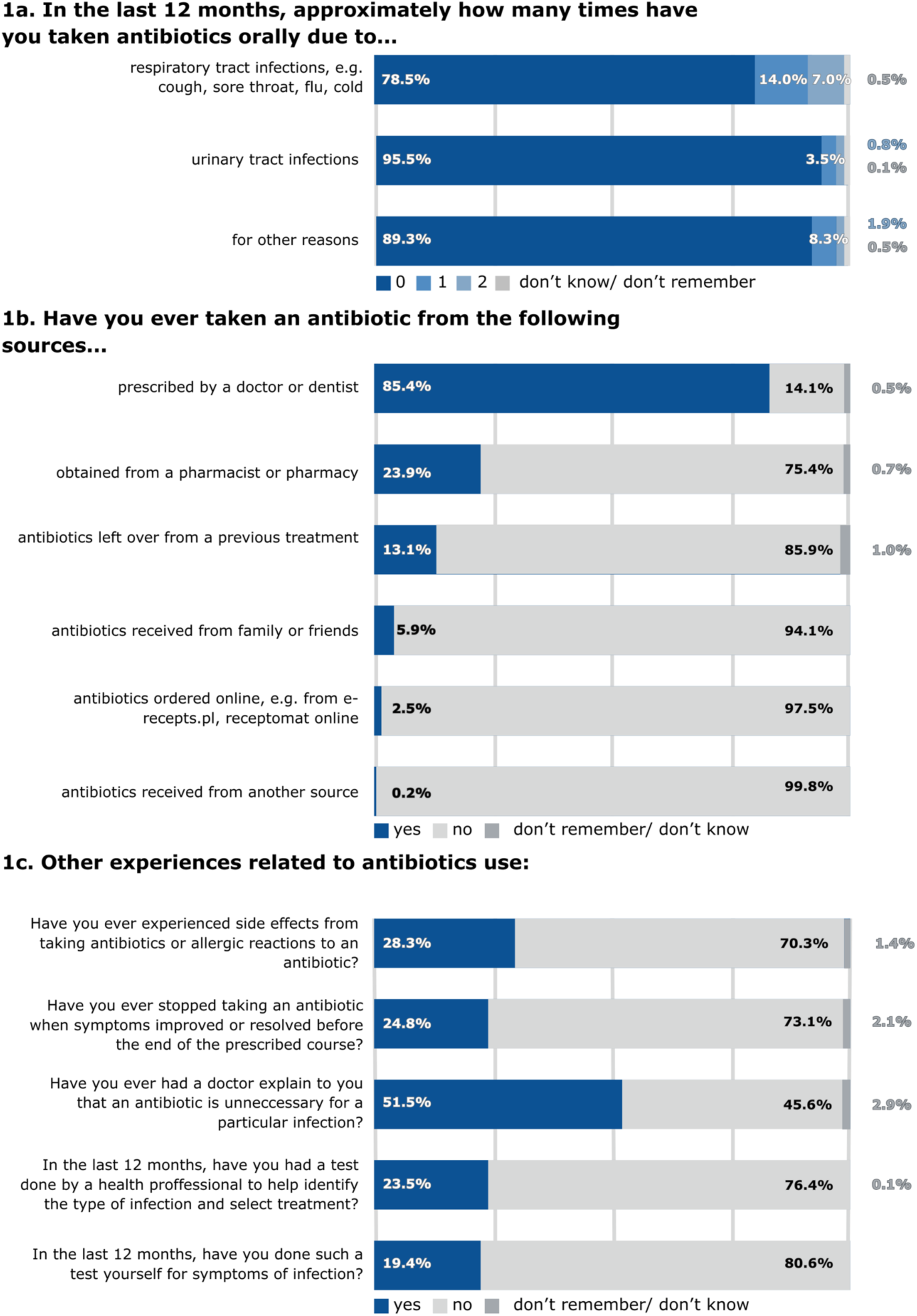
Self-reported experiences of infections and antibiotic-related behaviours. Survey results related to antibiotic use in the previous 12 months and other antibiotic-related behaviours and experiences (N=1,000)

A statistically significant effect of age, household income level (i.e. average household income per household member), and education level on antibiotic use (i.e. sum of all types of antibiotics taken in the preceding year) was observed. Older adults (aged 66+) and the middle-aged cohort (aged 36-65) had significantly greater odds of using antibiotics at least twice in the past year compared to younger adults (aged 18-35) (older adults: OR=8.14, 95% CI [2.44-27.14], p<0.001; middle-aged adults: OR=3.08, 95% CI [1.20-7.90], p=0.02). Respondents within the highest income bracket demonstrated an increased likelihood of using a single course of antibiotics in the past year, compared to those in the lowest income group (OR=3.24, 95% CI [1.45-7.25], p=0.004). Individuals with a vocational education (OR =3.62, 95% CI [1.20-10.91], p=0.02) or higher education (OR=3.80, 95% CI [1.04-13.83], p=0.04) had significantly greater odds of using antibiotics at least twice compared to those with the lowest level of education. No effect was found for respondents with secondary/post-secondary education. There was no significant effect of sex and settlement size on antibiotic use.

### Sources of antibiotics

The reported sources of antibiotics are presented in Figure 1b. Settlement size and age were associated with using leftover antibiotics. People living in large cities had significantly greater odds of using leftover antibiotics compared to those from rural areas (OR=1.81, 95% CI [1.04-3.16], p=0.04). Middle-aged adults were more likely than the youngest group to use leftover antibiotics (OR=2.05, 95% CI [1.19-3.56], p=0.04). This effect was not significant for older adults (p=0.58). Respondents who reported using leftover antibiotics were more likely to have taken antibiotics once in the preceding year (OR=2.47, 95% CI [1.36-4.48], p=0.003). Middle-aged (OR=0.51, 95% CI [0.27-0.96], p=0.04) and older adults (OR=0.29, 95% CI [0.11-0.75], p=0.01) had significantly lower odds of obtaining antibiotics from family and friends, compared to the youngest group. No significant effect of obtaining antibiotics from family and friends on antibiotic intake was observed (p≥0.60).

### Experiences of side effects

Among other experiences related to antibiotic use (Fig. 1c), 28.3% respondents reported experiencing side effects or allergic reactions when taking antibiotics. Women were more likely than men to report experiencing side effects or allergic reactions in the preceding year (OR=1.80, 95% CI [1.31-2.47], p<0.001). Moreover, middle-aged adults were more likely to report side effects or allergic reactions, compared to young adults (OR=1.52, 95% CI [1.03- 2.25], p=0.04), similarly to individuals with higher education as compared to those with the lowest education level (OR=1.90, 95% CI [1.01-3.56], p=0.046). Experiencing side effects or allergic reactions to antibiotics was associated with using one course of antibiotics (OR=1.76, 95% CI [1.10-2.82], p=0.02).

### Self-management

Stopping antibiotic courses when feeling better was reported by 24.8% of respondents (Fig. 1c). Older adults showed a decreased likelihood of interrupting antibiotics before the prescribed courses compared to the youngest group (OR=0.45, 95% CI [0.26-0.80], p=0.006). The practice of stopping antibiotics when feeling better was associated with taking antibiotics once (OR=1.79, 95% CI [1.10-2.90], p=0.02) and twice or more (OR=2.83, 95% CI [1.45- 5.52], p=0.002) in the preceding year. 51.5% of respondents recalled a doctor explaining to them that an antibiotic was unnecessary for a particular infection. Furthermore, 86.2% respondents reported usually using over-the-counter remedies for infection symptoms.

### Diagnostic testing

Among respondents, 23.5% recalled having a diagnostic test performed by a healthcare professional in the preceding year to identify their infection and guide treatment (Fig. 1c), with those in small towns and medium-sized cities significantly more likely to report a test compared to those in rural areas (small towns: OR=1.76, 95% CI [1.17-2.67], p=0.007; medium-sized cities: OR=1.80,, 95% CI [1.13-2.84], p=0.01). 19.4% of respondents reported self-testing for infection symptoms. Compared to young adults, older adults were significantly less likely to self-test (OR=0.45, 95% CI [0.25-0.81], p=0.008), as were individuals with secondary and post-secondary education compared to those with an elementary education (OR=0.36, 95% CI [0.19-0.65], p<0.001). There was a similar but non-significant trend for other levels of education (p≥0.24). No association between self-testing and antibiotic use was found (p≥0.09).

### Beliefs about antibiotic efficacy, expected recovery, and side effects

The perceived need for antibiotics in case of various infections is reported in Figure 2a. 51.9% of respondents agreed that antibiotics typically facilitate faster recovery from RTIs (Fig. 2b). Older adults were more likely to believe that antibiotics accelerate recovery from RTIs, compared to young adults (OR=1.64, 95% CI [1.01-2.65], p=0.045). Additionally, individuals with a vocational education had significantly greater odds of believing in faster recovery due to antibiotics than those with the lowest level of education (OR=1.74, 95% CI [1.07-2.85], p=0.03). A similar, though non-significant, trend was observed for other education levels (p≥0.06). People who believed that antibiotics facilitate faster recovery from RTIs were more likely to use antibiotics at least twice in the preceding year (OR=4.07, 95% CI [1.98-8.36], p<0.001). 48.3% of respondents expressed concern about the side effects of antibiotics (Fig. 2b). Individuals who reported experiencing side effects or allergic reactions to antibiotics were more likely to fear antibiotic side effects (OR=2.98, 95% CI [2.15-4.13], p<0.001). However, concerns about side effects were not associated with antibiotic use (p≥0.61).

**Figure 2.**
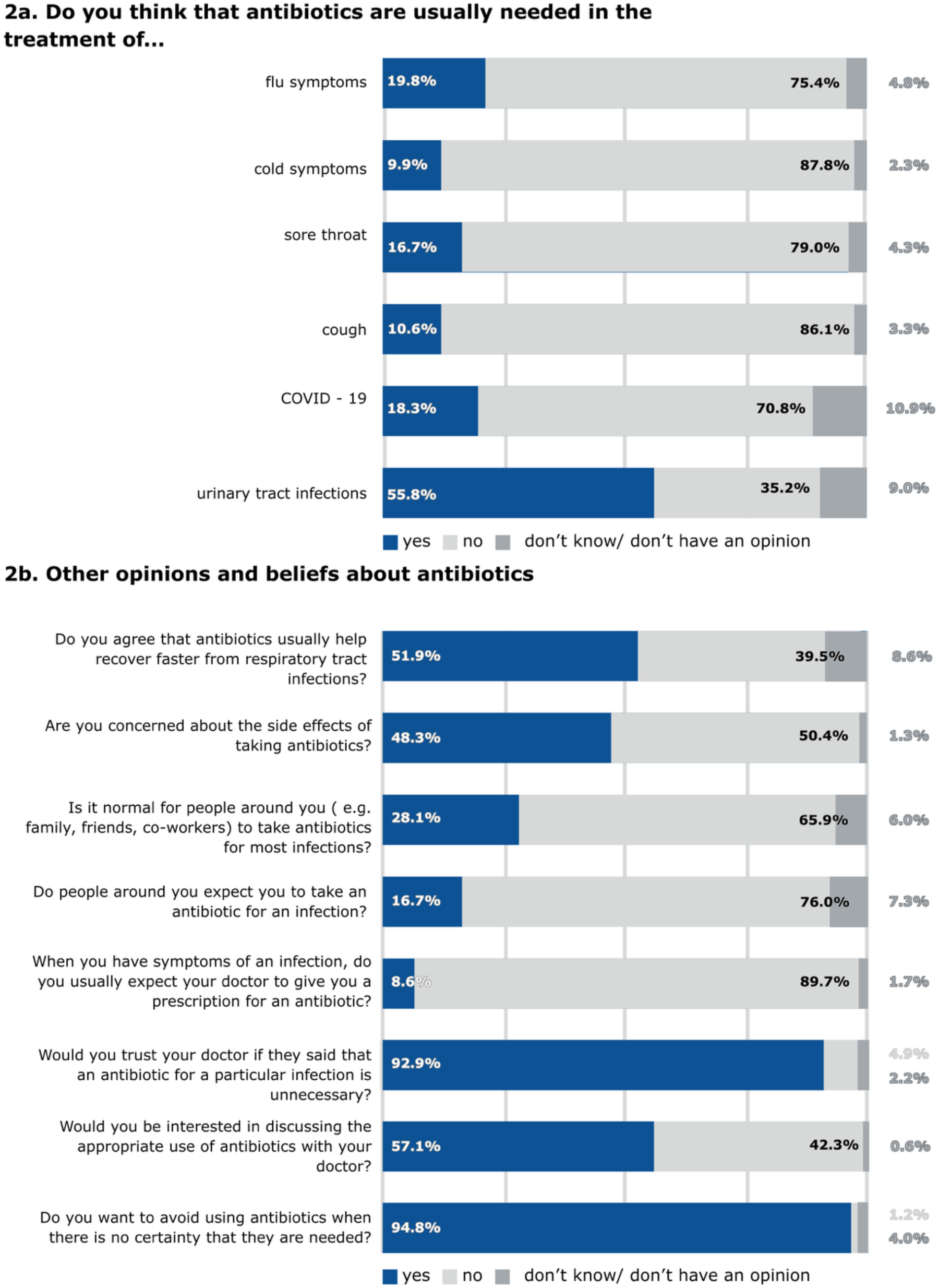
Self-reported beliefs about antibiotic use. Survey results related to beliefs about antibiotics (N=1,000)

**Figure 3.**
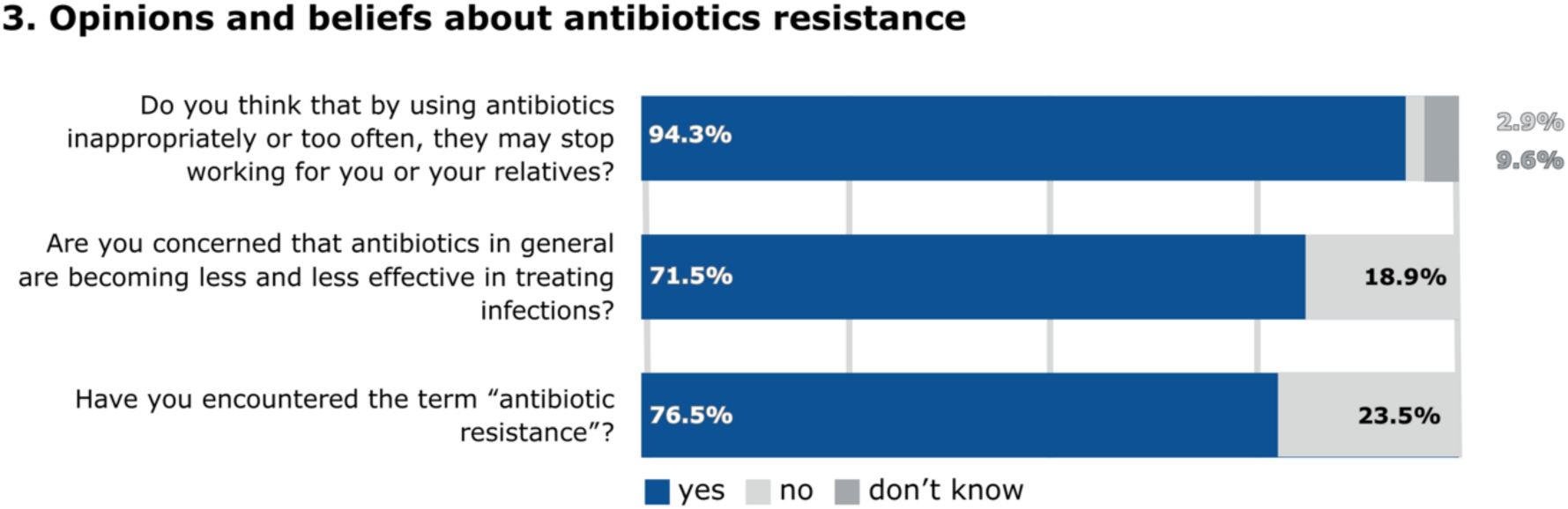
Self-reported beliefs about AMR. Survey results related to opinions and beliefs about antibiotic resistance (N=1,000)

### Social norms and social expectations

28.1% of respondents, especially older adults, compared to young adults (OR=1.67, 95% CI [1.01-2.76], p=0.04), considered it normal for people around them to take antibiotics for most infections (Fig. 2b). 16.7% of respondents felt pressured by others to take antibiotics for infections. Compared to men, women had significantly lower odds of reporting social expectations regarding antibiotic use (OR=0.67, 95% CI [0.46-0.97], p=0.04). Social norms and expectations were not associated with antibiotic use (p≥0.44).

### Expectations for antibiotics and the doctor-patient relationship

89.7% of individuals reported *not* typically expecting doctors to prescribe antibiotics when they have infection symptoms (Fig. 2b). However, older adults had significantly greater odds of expecting antibiotic prescriptions for infections compared to younger adults (OR=3.77, 95% CI [1.74-8.18], p<0.001). Expecting antibiotics was associated with the use of antibiotics at least twice in the past year (OR=3.47, 95% CI [1.39-8.69], p=0.008). 92.9% of respondents reported trusting their doctor if advised that antibiotics were unnecessary, and 57.1% expressed interest in discussing appropriate antibiotic use with their doctor. Individuals with a higher education were significantly less interested in discussing this matter with their doctor, compared to those with the lowest level of education (OR=0.57, 95% CI [0.32-0.997], p=0.049). A similar but non-significant trend was found for other education levels (p≥0.052). Expressing an interest in discussing appropriate antibiotic use with a doctor was associated with using antibiotics once (OR=2.05, 95% CI [1.31-3.21], p=0.002) and at least twice in the past year (OR=2.14, 95% CI [1.11-4.11], p=0.02). 94.8% of respondents expressed a wish to avoid antibiotics when their necessity was uncertain.

### Beliefs about AMR

94.3% of respondents believed that inappropriate or overuse of antibiotics could lead to antibiotics ceasing to be effective for themselves or their relatives. 71.5% expressed concern that antibiotics *in general* are becoming less effective in treating infections. Older adults were more likely to express this general concern than young adults (OR=2.27, 95% CI [1.24-4.16], p=0.008). Concerns about the effectiveness of antibiotics were not associated with antibiotic use (p≥0.17).

Furthermore, 76.5% of respondents reported having encountered the term “antibiotic resistance”. Individuals with a higher education were significantly more likely to report encountering this term than those with the lowest education level (OR=2.02, 95% CI [1.06- 3.87], p=0.03). A similar but non-significant trend was found for other education levels (p≥0.22).

When asked an open question about free associations with the term “antibiotic resistance”, 840 distinct responses were recorded from all respondents; 93.5% of respondents offered descriptions and 6.5% had no associations or did not know. The categories and frequencies of free-text responses are reported in Table 1. Most commonly, 44.1% of responses indicated a reasoning that antibiotic resistance refers to human bodies not reacting or being resistant to antibiotics. Other respondents described it in terms of antibiotic ineffectiveness (28.7%) and antibiotic misuse/overuse (20.5%). Only 12.8% respondents correctly defined antibiotic resistance as bacteria being/becoming resistant to antibiotics, antibiotics not (or no longer) working on bacteria or by giving examples of resistant (strains) of bacteria. A few respondents linked antibiotic resistance with microorganisms (in general or both bacteria and viruses), viruses, or “mutations”. Moreover, 11.8% of respondents described that antibiotic resistance means that antibiotics do not treat infections/illnesses or that illnesses are difficult/resistant to treatment, or mentioned viral illnesses or pneumonia.

**Table 1.**
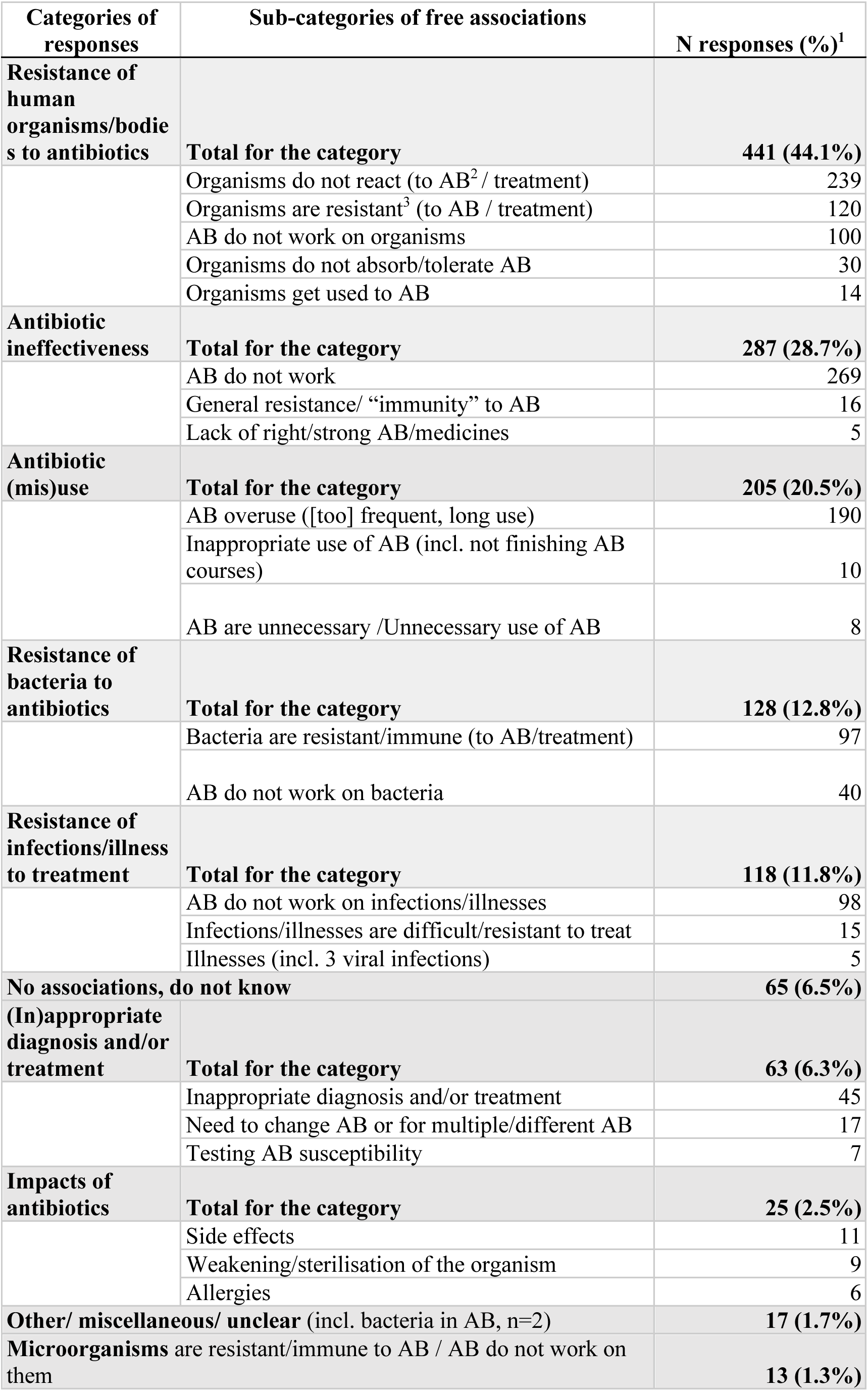
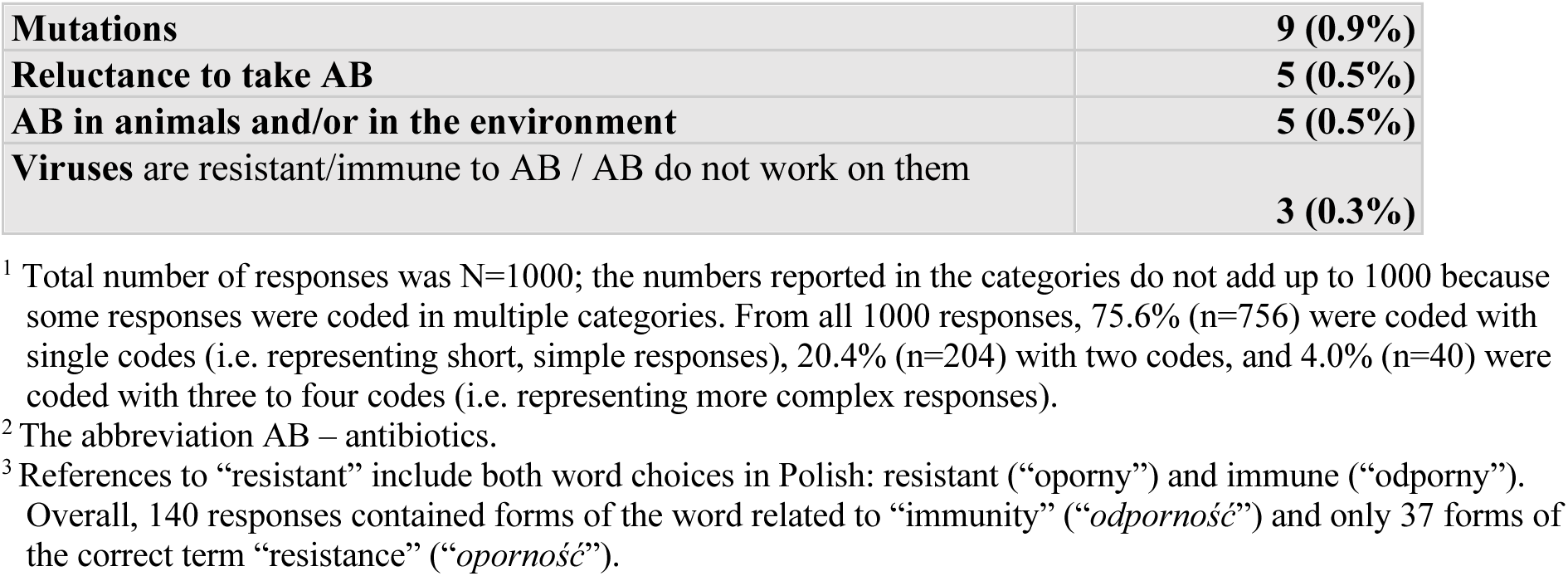
Types of responses on the free associations with the term “antibiotic resistance”.

| Categories of responses | Sub-categories of free associations | N responses (%)<sup>1</sup> |
| --- | --- | --- |
| **Resistance of human organisms/bodies to antibiotics** | **Total for the category** | **441 (44.1%)** |
|  | Organisms do not react (to AB<sup>2</sup> / treatment) | 239 |
|  | Organisms are resistant<sup>3</sup> (to AB / treatment) | 120 |
|  | AB do not work on organisms | 100 |
|  | Organisms do not absorb/tolerate AB | 30 |
|  | Organisms get used to AB | 14 |
| **Antibiotic ineffectiveness** | **Total for the category** | **287 (28.7%)** |
|  | AB do not work | 269 |
|  | General resistance/ “immunity” to AB | 16 |
|  | Lack of right/strong AB/medicines | 5 |
| **Antibiotic (mis)use** | **Total for the category** | **205 (20.5%)** |
|  | AB overuse ([too] frequent, long use) | 190 |
|  | Inappropriate use of AB (incl. not finishing AB courses) | 10 |
|  | AB are unnecessary /Unnecessary use of AB | 8 |
| **Resistance of bacteria to antibiotics** | **Total for the category** | **128 (12.8%)** |
|  | Bacteria are resistant/immune (to AB/treatment) | 97 |
|  | AB do not work on bacteria | 40 |
| **Resistance of infections/illness to treatment** | **Total for the category** | **118 (11.8%)** |
|  | AB do not work on infections/illnesses | 98 |
|  | Infections/illnesses are difficult/resistant to treat | 15 |
|  | Illnesses (incl. 3 viral infections) | 5 |
| **No associations, do not know** |  | **65 (6.5%)** |
| **(In)appropriate diagnosis and/or treatment** | **Total for the category** | **63 (6.3%)** |
|  | Inappropriate diagnosis and/or treatment | 45 |
|  | Need to change AB or for multiple/different AB | 17 |
|  | Testing AB susceptibility | 7 |
| **Impacts of antibiotics** | **Total for the category** | **25 (2.5%)** |
|  | Side effects | 11 |
|  | Weakening/sterilisation of the organism | 9 |
|  | Allergies | 6 |
| **Other/ miscellaneous/ unclear (incl. bacteria in AB, n=2)** |  | **17 (1.7%)** |
| **Microorganisms are resistant/immune to AB / AB do not work on them** |  | **13 (1.3%)** |

|  |  |
| --- | --- |
| **Mutations** | **9 (0.9%)** |
| **Reluctance to take AB** | **5 (0.5%)** |
| **AB in animals and/or in the environment** | **5 (0.5%)** |
| **Viruses are resistant/immune to AB / AB do not work on them** | **3 (0.3%)** |

<sup>1</sup> Total number of responses was N=1000; the numbers reported in the categories do not add up to 1000 because some responses were coded in multiple categories. From all 1000 responses, 75.6% (n=756) were coded with single codes (i.e. representing short, simple responses), 20.4% (n=204) with two codes, and 4.0% (n=40) were coded with three to four codes (i.e. representing more complex responses).
<sup>2</sup> The abbreviation AB – antibiotics.
<sup>3</sup> References to “resistant” include both word choices in Polish: resistant (“oporny”) and immune (“odporny”). Overall, 140 responses contained forms of the word related to “immunity” (“odporność”) and only 37 forms of the correct term “resistance” (“oporność”).

## Discussion

In our representative survey of the Polish general population, 31.6% reported using antibiotics in the preceding year, a proportion similar to that found between 2009-2011 (8) and almost double the rate reported in Poland in 2022, at the end of the COVID-19 pandemic (19).

Our findings demonstrate that sociodemographic factors – specifically higher age, income, and education level – play a significant role in predicting increased antibiotic use. Beyond demographics, the study highlights the importance of misconceptions regarding recovery speed and expectations for prescriptions. Although patients expressed high trust in their physicians, the data suggest that communication regarding the non-necessity of antibiotics remains suboptimal. Furthermore, 76.5% of respondents claimed to be aware of AMR, their qualitative descriptions frequently revealed a lack of genuine understanding, pointing to a gap between perceived and actual knowledge.

### Sociodemographic predictors of antibiotic use and related beliefs

We found that age, household income, and education level were significantly associated with antibiotic use. Sex and settlement size were not found to be significantly associated with antibiotic use. This contradicts prior research from Poland and across Europe, which showed that women used more antibiotics (9,19,31). While previous studies reported mixed results regarding the association between the settlement size and antibiotic-related behaviours (8,9,20,31,32), settlement size was not identified as a key factor in our study.

Age, in particular, was strongly associated with taking multiple courses of antibiotics. Older adults (over 66 years old) reported the highest antibiotic use, which may be attributed to a higher susceptibility to infections (33) and more frequent prescriptions (34). Nevertheless, we found that older adults held several beliefs that may potentially lead to increased antibiotic use: they were more likely to believe antibiotics accelerate recovery from RTIs, expect antibiotic prescriptions, and perceive antibiotic use as a social norm. Previous Polish research associated increasing age with more frequent antibiotic use, with middle-aged adults being the highest consumers (9). In our study, middle-aged adults (36-65 years old) used more antibiotics than younger adults but fewer than the oldest group. Middle-aged adults were more likely to use leftover antibiotics and were more aware of their side effects, suggesting a more self-directed approach than the oldest group.

Numerous European studies included higher education level as a significant predictor of good antibiotic knowledge (19,35,36). The relationship between education and the use of antibiotics is unclear, though our study identified education as a non-linear predictor of antibiotic use. Further research is needed to explore additional factors influencing this association.

Our results differ from general European trends, where lower-income groups were the most frequent antibiotic users (19). We found that individuals with the highest household income were most likely to use antibiotics, which is consistent with another study conducted in Poland (9). Higher antibiotic use among the high-income group might be linked to better access to private healthcare or to other, unknown factors. This study found no significant association between income and antibiotic beliefs or other related behaviours, and further research is needed to understand the association between income and antibiotic consumption.

### Psychosocial predictors of antibiotic use

Several misconceptions about antibiotics were identified. The most prominent in our study was the belief that antibiotics help recover faster from RTIs, despite most of the respondents believing that antibiotics are not usually needed for RTIs. Individuals believing in faster recovery from RTIs due to antibiotic treatment, who were typically older adults, were significantly more likely to have used antibiotics multiple times in the preceding year.

Our findings suggest that a high level of patient trust in doctors and willingness to discuss antibiotic use could be a promising foundation for future interventions, particularly as patients’ (mainly older adults’) expectations from doctors were associated with high antibiotic use. This is especially important given that the perceived patient pressure is among the most important factors influencing antibiotic prescribing in Poland (37). People trusted their doctors and had either discussed or were open to discussing antibiotic use with them, which contrasts previous findings in Poland showing that people are often dissatisfied with doctors’ advice (9) and frequently choose to self-medicate (10).

### Beliefs related to AMR

In line with findings from across the European Union (19), a majority (94.3%) of participants worried that antibiotics could become ineffective for themselves or their family due to inappropriate use, a concern also widely held on a more general level. However, a proper understanding of how antibiotics work and the nature of AMR was lacking throughout Europe and Poland (8,19,36). In our study, three-quarters of respondents reported having heard the term “antibiotic resistance”, but many of the free-text responses reflected a fundamental misunderstanding of the issue. The most common description was that a person’s body becomes resistant to antibiotics. Only 12.8% of participants accurately associated the term with bacteria becoming resistant to antibiotics or developing resistance. Confusing AMR with a whole-organism immunity, rather than antimicrobial resistance, is prevalent in many countries beyond Poland (38,39). This perception of AMR can lead to undesirable psychological consequences, including feelings of guilt or a diminished sense of control over one’s health. In the Polish context, this challenge is further complicated by linguistic similarities. Most respondents used the incorrect term “immunity” (“*odporność*”) to describe AMR, likely due to the linguistic similarity with the Polish word for “resistance” (“*oporność*”), which indicates a common and challenging public misconception that needs to be addressed in future communication campaigns.

### Implications for behaviour change interventions

Our findings offer implications for future interventions designed to promote prudent antibiotic use, especially through addressing the need for tailored interventions targeting high-risk groups like older adults. The recommendations are framed within the components of the COM-B model (18).

#### Correcting AMR misconceptions (Capability)

Public health campaigns should move beyond general awareness to actively correct public misunderstandings regarding AMR, enhancing AMR-related health literacy and psychological capability through clear, accurate education on transmission and treatment necessity.

#### Utilising the point-of-care interaction (Opportunity)

Intervention design should capitalise on the social opportunity within clinical settings. One-on-one conversations about prudent antibiotic use may be systematically integrated into routine practice, particularly with frequent antibiotic users.

#### Managing expectations for prescriptions and recovery (Motivation)

Interventions should directly challenge reflective motivation by promoting a realistic understanding of recovery timelines and reducing the high expectation of receiving an antibiotic prescription among older adults and frequent users.

### Strengths and limitations

The study’s primary strength is the use of a representative sample, which allows for the generalisation of findings to the Polish population. Our comprehensive analysis of both sociodemographic and psychosocial factors offers a strong foundation for future research and interventions aimed at optimising antibiotic use and mitigating AMR. The questionnaire was systematically developed and included novel questions on psychosocial aspects of antibiotic use. The use of an open-ended question on AMR also provided a more nuanced understanding of public perceptions. Key limitations include the potential exclusion of certain groups (e.g., minorities without mobile phones) and no data on reasons for declining to participate or withdrawing from the survey. Another limitation is the reliance on respondents being able to recall their antibiotic use and knowledge of which medications are antibiotics, which might have led to potential underreporting. It is possible that social desirability bias affected the responses from participants, even though we aimed to minimise this by phrasing our questions in a neutral tone and ensuring anonymity. Additionally, we were unable to collect information on respondents’ health status, which limited our ability to link antibiotic use to specific clinical factors, as the CATI method - while crucial for accessing a representative sample - is generally unsuitable for highly personal inquiries and lacks the capacity required for the precise measurement of health status using standardised clinical scales and supplementary medical data.

### Conclusions

This study described antibiotic-related behaviours, experiences, and beliefs of the general population in Poland. Our primary finding is that adults older than 66 years are characterised by frequent antibiotic use, high prescription expectations, and perceiving antibiotic use as a social norm, and thus may be considered a priority population group for targeted interventions. Health promotion and health educations interventions and campaigns should simultaneously help manage patient expectations regarding recovery timelines and prescriptions. A key opportunity exists in leveraging clinical interactions for personalised education, especially with frequent antibiotic users who show a willingness to discuss antibiotics with physicians. Finally, public health efforts must transcend superficial awareness of AMR and actively focus on correcting deep-seated public misconceptions to drive meaningful behavioural change.

## Data Availability

The datasets generated during the current study will be available in the SWPS Science Share repository.

## Abbreviations

AMR: antimicrobial resistance
RTI: respiratory tract infection
UTI: urinary tract infection
EU: European Union

## Acknowledgements

Not applicable

## Declarations

### Ethics approval and consent to participate

Ethics approval reference: 02/E/04/2025, reviewed and approved by Department of Psychology Ethics Committee in Wrocław, SWPS University. The study was conducted in accordance with the ethical principles outlined Declaration of Helsinki. No identifiable data were collected from respondents, and all participants provided informed consent prior to their inclusion in the study, having been apprised of the research objectives and procedures.

Participation was voluntary, and respondents could withdraw at any time without consequence.

### Consent for publication

Not applicable

## Supplementary Materials include

STROBE checklist, study questionnaire (Polish and English version), study participants characteristics, distribution of responses to questionnaire questions, results of logistic regression analysis.

